# Attention-Deficit Hyperactivity Disorder and Intimate Partner Violence Perpetration: Sex Differences and Subtype-Specific Associations in a Population-Based Study

**DOI:** 10.1101/2025.10.08.25337602

**Authors:** Yuting Wu, Kia-Chong Chua, Sally McManus, Zoe Morgan, Sian Oram

## Abstract

**Background:** Attention-deficit hyperactivity disorder (ADHD) has been linked to interpersonal violence, but its relationship with intimate partner violence (IPV) perpetration, particularly in women and across symptom subtypes, remains underexplored. This study examines associations between ADHD and IPV perpetration in a nationally representative sample, with a focus on sex differences and ADHD symptom dimensions.

**Methods:** Data were drawn from the 2014 Adult Psychiatric Morbidity Survey (APMS), a cross-sectional household survey of adults aged 16+ in England. Positive screen for ADHD was assessed using the 6-item Adult ADHD Self-Report Scale (ASRS), with subscale scores for inattentiveness and hyperactivity/impulsivity calculated. IPV perpetration was measured via four self-completed items capturing psychological, physical, and sexual violence. The inclusion criteria for the analysis were adults who had been in a relationship. Logistic regression models estimated associations between ADHD and IPV perpetration, adjusting sequentially for sociodemographic, trauma, behavioural, and psychiatric covariates. Marginal effects of interaction terms between sex and ADHD symptoms were used to model predicted probabilities across symptom severity.

**Results:** ADHD was associated with increased odds of IPV perpetration (OR 2.70, 95% CI: 2.12-3.44). This association attenuated after adjustment for covariates. Inattentiveness remained associated with IPV perpetration after full adjustment; hyperactivity/impulsivity was marginal. The ADHD-IPV relationship differed by sex: across all ADHD subtypes, predicted probabilities of IPV perpetration increased with symptom severity in women but not in men.

**Conclusions:** ADHD may contribute to risk of IPV perpetration, with stronger effects among women. These findings highlight the importance of dimensional ADHD assessments, sex-specific analyses, and tailored intervention approaches for IPV prevention among individuals with neurodevelopmental and mental health needs.

## Background

Intimate partner violence (IPV) is a significant public health concern, encompassing physical, sexual, and psychological violence perpetrated by a current or former romantic partner (World Health Organization, 2012). Globally, more than one in four ever-partnered women aged 15-49 years report having experienced IPV (World Health Organization, 2021). Victimisation has been linked to increased risks of mental disorders, including depression, anxiety, and post-traumatic stress disorder (Oram et al., 2022; White et al., 2024)

IPV perpetration is a heterogeneous phenomenon, driven by an interplay of individual-level risk factors and broader contextual influences (Gibbs et al., 2020). Although people with mental disorders are more likely to be victims than perpetrators of violence, mental disorders are associated with an elevated risk of IPV perpetration (Oram et al., 2014; Yu et al., 2019). Across all mental disorders, substance use disorders exhibit the strongest and most consistent link to IPV perpetration, with comorbid substance misuse amplifying risk across diagnostic groups.

Emerging evidence suggests that attention-deficit hyperactivity disorder (ADHD) may be associated with an increased risk of IPV perpetration (Arrondo et al., 2023; Buitelaar et al., 2020). ADHD is characterised by impairments in cognitive, emotional, and executive functioning, particularly impulsivity, hyperactivity, and difficulties with self-regulation, traits also commonly observed in individuals who engage in aggressive behaviour (Biederman et al., 2004; Christiane Desman et al., 2008; Retz et al., 2021; Roselló et al., 2020; Soler-Gutiérrez et al., 2023). Additionally, adults with ADHD have higher rates of comorbid mental disorders, including substance use disorders, depressive disorders, bipolar disorders, and personality disorders, (Choi et al., 2022; Mak et al., 2020; Mak et al., 2022), all of which have been associated with an increased risk of IPV perpetration (Abramsky et al., 2011; Gibbs et al., 2020; Oram et al., 2022; Spencer et al., 2019).

A 2023 systematic review and meta-analysis synthesising findings from seven studies identified an association between ADHD and IPV perpetration, though effect sizes varied and the role of potential confounders, such as substance misuse and antisocial personality disorders, remained unclear (Arrondo et al., 2023). A population-based study using data from the 2007 Adult Psychiatric Morbidity Survey (APMS) found that after adjusting for comorbidities among individuals who screened positive for ADHD, hyperactivity/impulsivity remained significantly associated with having been in a physical fight with, or having deliberately hit, a partner while inattentiveness did not (Gonzalez et al., 2013). This finding suggested that impulsivity, rather than inattention, may be the primary ADHD-related contributor to IPV risk, aligning with findings of a strong association between hyperactivity/impulsivity and violent behaviour or executive dysfunction (Arrondo et al., 2023; C. Desman et al., 2008).

Most studies have focused on male samples. A review by Buitelaar et al. (2020) found that three out of four case-control studies on ADHD and IPV examined only males. It is therefore unclear whether the association between ADHD and IPV perpetration is specific to males, or whether it extends to females. Research on mental disorder and IPV has identified sex differences in mental health-related IPV perpetration risks. While depression has been associated with IPV perpetration in women, for example, similar evidence for men remains scarce (Oram et al., 2014). In general, mental disorder has shown a more robust association with physical IPV perpetration among women, whereas evidence among men has been less consistent and predominantly based on select populations, such as military veterans with PTSD (Gibbs et al., 2020; Oram et al., 2022).

ADHD itself exhibits notable sex differences. Males with ADHD are more likely to present with hyperactive/impulsive symptoms, while females more commonly display inattentive symptoms (Rucklidge, 2010). Given that impulsivity and poor self-control have been strongly associated with violent behaviour (Arrondo et al., 2023), it is plausible that the relationship between ADHD and IPV perpetration varies by sex.

A recent study in Germany observed a consistent association between ADHD and both IPV perpetration and victimisation across genders and age groups (Merscher et al., 2025). While this adds to the growing recognition of ADHD as a risk factor for IPV, the study’s relatively modest sample size, non-probabilistic self-selected sampling strategy, and reliance on an online self-report survey underscore the need for further investigation into sex-specific associations using representative samples and dimensional assessments of ADHD symptoms.

This study therefore aimed to examine the relationship between screening positive for ADHD and lifetime IPV perpetration, with a focus on sex differences and ADHD symptom subtypes. Specific objectives were to: (1) estimate the association between ADHD and lifetime IPV perpetration in a population-based sample; (2) investigate sex differences in the relationship between ADHD and IPV perpetration; (3) assess ADHD subtypes (inattentiveness vs. hyperactivity/impulsivity) in relation to IPV perpetration and explore whether these associations differ by sex.

## Methods

### Study design and data source

We analysed data from the 2014 Adult Psychiatric Morbidity Survey (APMS), a cross-sectional survey that is nationally representative of the household population of England (aged 16 years and older). The survey uses stratified, multi-stage random sampling approach based on the Small User Postcode Address File. Full details of sampling are reported elsewhere (McManus, Bebbington, et al., 2019). Data were collected between May 2014 and September 2015, with a final sample of 7,546 participants (57% response rate).

Interviews were conducted in people’s homes by trained interviewers using a combination of computer-assisted interviewing and, for some sensitive questions, computer-assisted self-completion interviews. The 2014 APMS was approved by the West London National Research Ethics Committee (reference number: 14/LO/0411). Ethical approval for this analysis was provided by the LSHTM Research Ethics Committee (reference number: 31013).

### Measures

#### ADHD

ADHD was screened for using the 6-item Adult ADHD Self-Report Scale (ASRS) (Kessler et al., 2007). The scale assesses the frequency of symptoms over the past six months on a five-point Likert scale (ranging from 0, ‘never’, to 4, ‘very often’). Each item was dichotomized based on established threshold frequencies, and a total score was derived by summing the responses. A total score of four or more was considered indicative of a ‘positive screen for ADHD,’ resulting in a binary variable reflecting the presence or absence of ADHD.

Four items within the 6-item ASRS assess inattentiveness and two evaluate hyperactivity/impulsivity (Table 1). Continuous variables representing inattentiveness and hyperactivity/impulsivity were generated by summing the respective item scores, with maximum possible scores of 16 and 8, respectively. In addition to the standard 6-item ASRS, the 2014 APMS included an additional item from the full 18-item ASRS to further assess hyperactivity/impulsivity (’how often do you have difficulty waiting your turn in situations where turn-taking is required?’). Although this item was not included in the overall ADHD score, it was considered relevant for capturing a specific aspect of hyperactivity/impulsivity. Consequently, we also constructed a three-item measure of hyperactivity/impulsivity with a maximum score of 12.

**Table 1.**
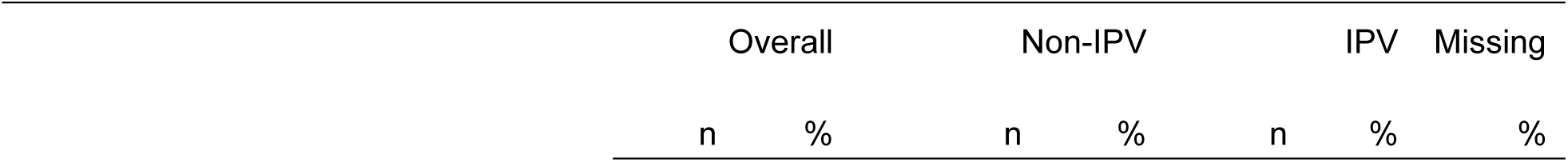
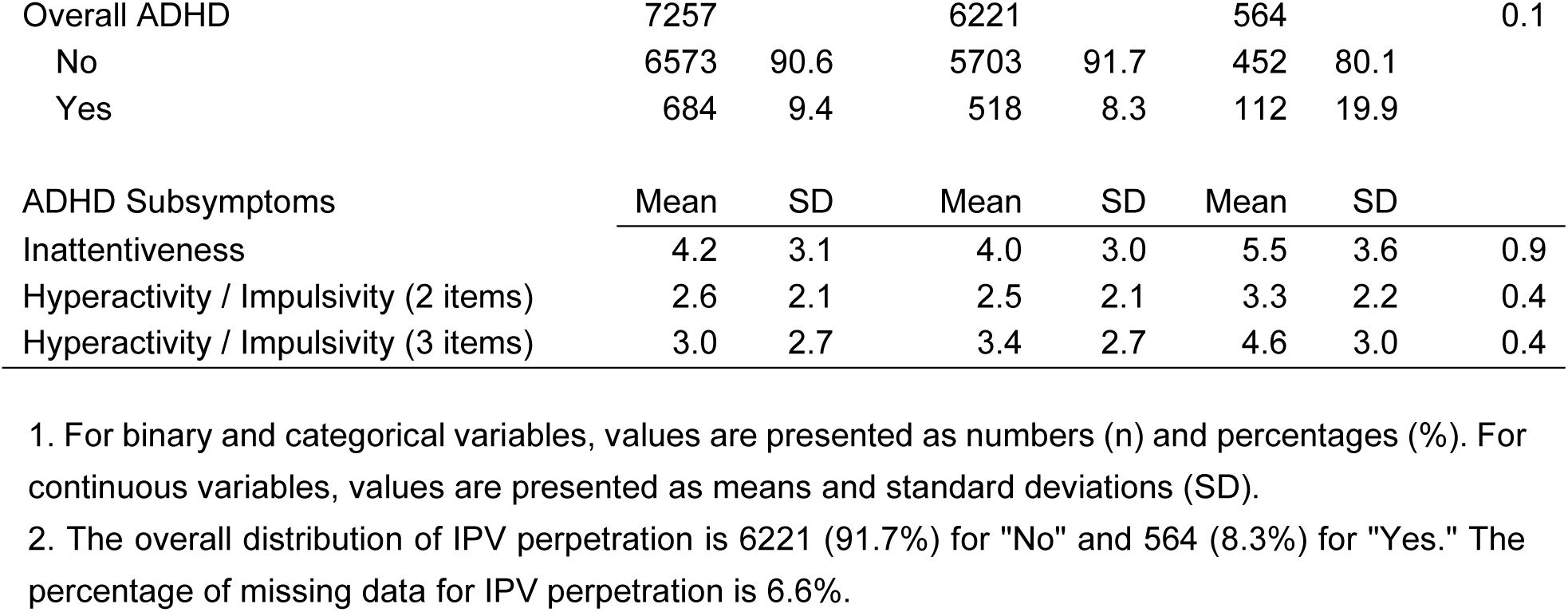
Distribution of ADHD Exposures by IPV perpetration.

| Overall |  | Non-IPV |  | IPV |  | Missing |
| --- | --- | --- | --- | --- | --- | --- |
| n | % | n | % | n | % | % |

|  |  |  |  |  |  |  |  |
| --- | --- | --- | --- | --- | --- | --- | --- |
| Overall ADHD | 7257 |  | 6221 |  | 564 |  | 0.1 |
| No | 6573 | 90.6 | 5703 | 91.7 | 452 | 80.1 |  |
| Yes | 684 | 9.4 | 518 | 8.3 | 112 | 19.9 |  |
| ADHD Subsymptoms | Mean | SD | Mean | SD | Mean | SD |  |
| Inattentiveness | 4.2 | 3.1 | 4.0 | 3.0 | 5.5 | 3.6 | 0.9 |
| Hyperactivity / Impulsivity (2 items) | 2.6 | 2.1 | 2.5 | 2.1 | 3.3 | 2.2 | 0.4 |
| Hyperactivity / Impulsivity (3 items) | 3.0 | 2.7 | 3.4 | 2.7 | 4.6 | 3.0 | 0.4 |

1. For binary and categorical variables, values are presented as numbers (n) and percentages (%). For continuous variables, values are presented as means and standard deviations (SD).
2. The overall distribution of IPV perpetration is 6221 (91.7%) for "No" and 564 (8.3%) for "Yes." The percentage of missing data for IPV perpetration is 6.6%.

#### IPV perpetration

Lifetime IPV perpetration was measured through four questions adapted from the Conflict Tactics Scale (Straus, 1979). Questions formed part of the self-completion module for respondents who reported having ever been in a relationship. One question measured psychological IPV, two measured physical IPV, and one measured sexual violence. Each was coded as a binary variable. We generated a composite binary variable for any lifetime IPV perpetration where responding yes to one or more questions was coded as a positive response.

#### Covariates

Covariates of interest included antisocial and borderline personality disorder, substance use, perpetration of non-partner violence, IPV victimisation, childhood abuse and neglect, and use of ADHD medication. Antisocial and borderline personality disorders are frequently comorbid with ADHD and have been independently linked to IPV perpetration in both men and women (Spencer et al., 2019; Storebo & Simonsen, 2016). Substance use disorders are highly prevalent among individuals with ADHD (Kessler et al., 2006; Wilens et al., 2011) and are established risk factors for IPV perpetration, partly due to their effects on cognitive disinhibition and aggression (Cafferky et al., 2018; Foran & O’Leary, 2008; Steele & Josephs, 1990). Perpetration of non-partner violence captures generalised aggression, which often co-occurs with IPV perpetration and may reflect broader patterns of violent behaviour beyond intimate relationships (Bernat et al., 2012; Theobald & Farrington, 2012). IPV victimisation is also a relevant covariate, particularly given evidence that mutual violence is common and that victimisation may precede or occur alongside perpetration, especially in the context of self-defensive aggression (Holmes et al., 2022; Palmetto et al., 2013; Wymbs et al., 2019). Childhood abuse and neglect are associated both with ADHD symptomatology (Aguado-Gracia et al., 2021; Ouyang et al., 2008) and with increased risk of later IPV perpetration, possibly via their long-term effects on emotion regulation and interpersonal functioning (Brennan et al., 2021; Widom et al., 2014). Finally, the use of ADHD medication may attenuate associations between ADHD and these risk factors, given evidence that pharmacological treatment is linked to reduced criminal behaviour and improved cognitive functioning (Lichtenstein et al., 2012; Ruiz-Goikoetxea et al., 2018). Demographic and socioeconomic variables were included as covariates because both ADHD and IPV perpetration have been linked to specific social and structural factors. Individuals with ADHD are more likely to be male, unemployed, and previously married (Kessler et al., 2006), while IPV perpetration has been associated with lower socioeconomic status, including unemployment, low income, and younger age (Caetano et al., 2008; Stith et al., 2004; Wilson, 2019). Including these variables helps account for potential confounding arising from shared sociodemographic risk profiles.

#### Personality disorder

ASPD and BPD were assessed using the self-completion Structured Clinical Interview for DSM-IV Personality Disorders (SCID-II) (First et al., 1997), administered in a self-completion section among participants aged 16-64 years. We created a binary variable for personality disorder based on presence of either ASPD or BPD.

#### Substance use

Substance use was assessed using self-completion data on hazardous alcohol use and drug dependence. Hazardous alcohol use was defined as scoring ≥8 on the Alcohol Use Disorders Identification Test (AUDIT), with dependence further assessed using the Severity of Alcohol Dependence Questionnaire (SADQ), where scores ≥4 indicated dependence. Signs of drug dependence were assessed for 8 commonly used substances, based on past-year use and self-reported symptoms. A binary variable was created indicating substance use if either hazardous alcohol use or drug dependence was present.

#### Perpetration of non-partner violence

Perpetration of non-partner violence was assessed using an ASPD screening item asking whether participants had engaged in a physical fight, assault, or deliberately hit someone in the past five years. Those who responded “Yes” were asked to identify the individuals involved; participants who selected any category other than a spouse or partner (e.g., child, family member, friend, stranger) were coded as having perpetrated non-partner violence. This was recorded as a binary variable, with limited overlap observed with ASPD diagnoses.

#### IPV victimisation

IPV victimisation was measured using five binary (Yes/No) self-completion items covering physical (three items), economic (one item), and psychological or emotional abuse (one item). Participants who answered “Yes” to at least one item were classified as having experienced IPV victimisation. This was coded as a binary variable indicating any lifetime experience of IPV.

#### Childhood neglect and abuse

Childhood abuse and neglect were assessed via 14 self-completion items covering neglect (7 items on a 5-point frequency scale), physical abuse (4 binary items), and sexual abuse (3 binary items). Neglect responses were recoded so that any answer other than “Never” was coded as “Yes.” To avoid loss of statistical power due to high prevalence, responses across all items were summed to create a continuous variable reflecting the total number of types of childhood abuse and neglect experienced.

#### ADHD medication

ADHD medication use was assessed during face-to-face interviews, where participants were asked whether they had ever taken any listed psychiatric medications. A binary variable was created to indicate use of ADHD medication, with “Yes” assigned to those who reported currently taking either Ritalin or Strattera.

#### Sociodemographic factors

Demographic and socioeconomic covariates included age (continuous), sex (male/female), ethnicity (White/Minority groups, including Black, Asian, and Mixed), marital status (partnered, including married, cohabiting, and same-sex couples / non-partnered, including single, widowed, divorced, and separated), and employment status (employed / non-employed, combining unemployed and economically inactive). Dichotomisation of ethnicity, marital status, and employment addressed small cell sizes and improved model stability and interpretability.

### Statistical Analysis

All analyses accounted for the complex survey design of the 2014 APMS, using population weights to adjust for unequal selection probabilities, non-response, and post-stratification based on age, sex, and region using mid-year ONS population estimates. The dataset was weighted using the syvset command in STATA/SE18.0.

Descriptive statistics were calculated for all variables of interest, with continuous variables summarised as means and standard deviations, and categorical variables as frequencies and percentages. Analyses excluded participants who reported never having been in an intimate relationship.

The association between ADHD and IPV perpetration was examined using a series of logistic regression models, following Gonzalez et al. (2013). Model 1 adjusted for sociodemographic factors (including age, sex, ethnicity, marital status, and employment status). Model 2 additionally adjusted for life events (childhood abuse and neglect, and IPV victimisation), and Model 3 further adjusted for behavioural and mental health factors (substance use, personality disorder, and non-partner violence perpetration). To explore potential moderation by sex, Model 4 introduced an interaction term between ADHD and sex to the fully adjusted model (Model 3), followed by post-estimation of marginal effects to illustrate the predicted probability of IPV perpetration across ADHD status for men and women (Norton et al., 2019). To investigate potential mechanisms, the models were repeated using ADHD subtypes (inattentiveness and hyperactivity/impulsivity).

We conducted two sensitivity analyses. To assess whether differences in measurement scales introduced substantive variation in the association with IPV perpetration and its interaction with sex, the first sensitivity analysis used the 3-item version of the hyperactivity/impulsivity scale (in place of the 2-item version used in the primary models). Given that ADHD is a highly heritable condition that typically emerges early in life (Law et al., 2014; Sibley et al., 2022), and that longitudinal research suggests it increases vulnerability to subsequent trauma exposure, externalising behaviours, and psychiatric comorbidity (Posner et al., 2020; Storebø & Simonsen, 2016; Wilens et al., 2011), several covariates included in Models 2 and 3 may plausibly lie on the causal pathway between ADHD and IPV perpetration. To mitigate potential overadjustment and assess its influence on our main findings (Schisterman et al., 2009), we conducted a second sensitivity analysis (Model 5), in which the sex-by-ADHD interaction term was included in a model adjusting only for sociodemographic covariates (Model 1).

## Results

After excluding 201 individuals who reported having never been in a relationship, the sample size for analysis was 7,345. The sample was 51.8% male and 88.2% white, with a mean age of 47.8 years (SD 18.8). The majority were currently married or cohabiting (63.7%) and employed (59.9%). Missing data exceeded 5% for the following variables: lifetime IPV perpetration (6.6%), IPV victimization (5.8%), childhood abuse and neglect (5.8%), perpetration of non-partner violence (25.6%), and personality disorder (25.3%).

The prevalence of screen positive for ADHD was 9.4%, and the prevalence of reported IPV perpetration was 8.3%. As shown in Table 1, the prevalence of ADHD was higher among participants who reported IPV perpetration compared to those who did not (19.9% vs. 8.3%).

As shown in Table 2, perpetration of non-partner violence, personality disorder, substance use, and IPV victimization were higher among individuals with ADHD compared to those without ADHD and among IPV perpetrators compared to non-IPV perpetrators. Mean scores for childhood abuse and neglect are also higher in the ADHD group and among IPV perpetrators. Few participants reported ADHD medication use (n=6); this variable was excluded from subsequent analyses.

**Table 2.** Weighted Estimates of Sociodemographic Factors, Life Events, and Behavioural and Mental Health Factors by ADHD and IPV Perpetration Status.

|  |  | Overall | ADHD (%) |  | IPV (%) |  |
| --- | --- | --- | --- | --- | --- | --- |
|  | n | % | No | Yes | No | Yes |
| **Sociodemographic Factors** |  |  |  |  |  |  |
| Ethnicity | 7237 |  |  |  |  |  |
| White | 6380 | 88.2 | 88.3 | 87.4 | 88.6 | 91.1 |
| Black | 225 | 3.1 | 3.0 | 4.4 | 3.0 | 0.4 |
| Asian | 447 | 6.2 | 6.1 | 6.4 | 6.2 | 0.2 |
| Mixed | 185 | 2.6 | 2.6 | 1.8 | 2.3 | 2.2 |
| Sex | 7261 |  |  |  |  |  |
| Male | 3497 | 48.2 | 48.0 | 49.6 | 48.4 | 46.6 |
| Female | 3764 | 51.8 | 52.0 | 50.4 | 51.6 | 53.4 |
| Age | 7261 |  |  |  |  |  |
| Mean |  | 47.8 | 48.6 | 41.4 | 47.6 | 44.8 |
| (SD) |  | 18.8 | 18.9 | 16.3 | 18.7 | 15.6 |
| Marital status | 7261 |  |  |  |  |  |
| Partnered | 4627 | 63.7 | 65 | 51.2 | 65.1 | 58.1 |
| Non-partnered | 2634 | 36.3 | 35 | 48.8 | 34.9 | 41.9 |
| Employment status | 7261 |  |  |  |  |  |
| Employed | 4348 | 59.9 | 60.9 | 50.1 | 61.0 | 66.2 |
| Non-employed | 2913 | 40.1 | 39.1 | 49.9 | 39.0 | 33.8 |
| **Life Events** |  |  |  |  |  |  |
| Intimate partner violence victimization | 6839 |  |  |  |  |  |
| No | 5343 | 78.1 | 80.1 | 58.6 | 82.7 | 26.4 |
| Yes | 1497 | 21.9 | 19.9 | 41.4 | 17.3 | 73.6 |
| Childhood abuse and neglect | 6838 |  |  |  |  |  |
| Mean |  | 1.9 | 1.8 | 2.8 | 1.8 | 3.3 |
| (SD) |  | 2.4 | 2.9 | 2.3 | 2.3 | 3.1 |
| **Behavioural and Mental Health Disorders** |  |  |  |  |  |  |
| Perpetration of non-partner violence | 5401 |  |  |  |  |  |
| No | 4979 | 92.2 | 93.2 | 84.0 | 93.8 | 79.0 |
| Yes | 422 | 7.8 | 6.8 | 16.0 | 6.3 | 21.0 |
| Personality disorder | 5421 |  |  |  |  |  |
| No | 5149 | 95.0 | 96.2 | 84.9 | 96.5 | 81.1 |
| Yes | 272 | 5.0 | 3.8 | 15.1 | 3.5 | 18.9 |
| Substance use | 6994 |  |  |  |  |  |
| No | 5507 | 78.7 | 80.0 | 66.7 | 79.9 | 60.4 |
| Yes | 1487 | 21.3 | 20.0 | 33.3 | 20.1 | 39.6 |
| **Other** |  |  |  |  |  |  |
| ADHD medication | 7259 |  |  |  |  |  |
| No | 7253 | 99.9 | 99.9 | 99.6 | 100.0 | 99.6 |
| Yes | 6 | 0.1 | 0.1 | 0.4 | 0.0 | 0.4 |

1. Binary and categorical variables are presented as numbers (n) and percentages (%), while continuous variables (including age and childhood abuse and neglect) are shown as means and standard deviations (SD). 2. Ethnicity is shown in original categories to provide descriptive detail; however, a binary variable (White/Minority groups) was used in subsequent analyses to improve model stability. 3. Substance use was originally categorized as: "No", "Drug dependence / Hazardous alcohol use but no dependency / Alcohol dependence".

Table 3 presents the results of the logistic regression analyses examining the association between ADHD and IPV perpetration. In model 1, which adjusts for sociodemographic factors only, ADHD is significantly associated with increased odds of IPV perpetration (OR:2.70, 95% CI: 2.12–3.44). This association is attenuated after adjusting for adverse life events in model 2 (OR: 1.54, 95% CI: 1.14–2.08) and further reduced in model 3 with additional adjustment for violent behaviour and mental health disorders (OR 1.28, 95% CI: 0.92–1.77), rendering the association non-significant.

**Table 3.** Results of Logistic Regression for ADHD and IPV Perpetration from Models 1 to 4.

|  |  | Odds ratio | 95% CI (LB) | 95% CI (UB) | p-value |
| --- | --- | --- | --- | --- | --- |
| Model 1 | Overall ADHD | 2.70 | 2.12 | 3.44 | < 0.001 |
|  | Inattentiveness | 1.15 | 1.12 | 1.18 | < 0.001 |
|  | Hyperactivity / impulsivity | 1.17 | 1.12 | 1.23 | < 0.001 |
|  | Hyperactivity / impulsivity * | 1.15 | 1.11 | 1.20 | < 0.001 |
| Model 2 | Overall ADHD | 1.54 | 1.14 | 2.08 | 0.005 |
|  | Inattentiveness | 1.08 | 1.04 | 1.11 | < 0.001 |
|  | Hyperactivity / impulsivity | 1.08 | 1.02 | 1.14 | 0.006 |
|  | Hyperactivity / impulsivity * | 1.09 | 1.04 | 1.13 | < 0.001 |
| Model 3 | Overall ADHD | 1.28 | 0.92 | 1.77 | 0.144 |
|  | Inattentiveness | 1.05 | 1.01 | 1.09 | 0.012 |
|  | Hyperactivity / impulsivity | 1.03 | 0.97 | 1.10 | 0.353 |
|  | Hyperactivity / impulsivity * | 1.05 | 1.00 | 1.10 | 0.048 |
| Model 4 | Overall ADHD | 3.44 | 1.82 | 6.50 | < 0.001 |
|  | Inattentiveness | 1.09 | 1.02 | 1.17 | 0.014 |
|  | Hyperactivity / impulsivity | 1.20 | 1.07 | 1.34 | 0.001 |
|  | Hyperactivity / impulsivity * | 1.16 | 1.06 | 1.26 | 0.001 |

1. Hyperactivity /impulsivity \*: 3-item measure for sensitivity analysis.
2. LB and UB refer to the lower and upper bounds of the 95% confidence intervals, respectively.
3. Estimates that are not statistically significant or marginally significant are presented in grey.
4. Model 1 controls for sociodemographic factors.
Model 2 additionally controls for life events.
Model 3 further controls for other behavioural and mental health disorders.
Model 4 includes an interaction term between sex and ADHD.

A similar attenuation pattern is observed for the ADHD subtypes. Both inattentiveness and hyperactivity/impulsivity show significant associations with IPV perpetration in model 1, which progressively diminish across subsequent models. In model 3, the association between hyperactivity/impulsivity and IPV perpetration was no longer statistically significant (OR: 1.03, 95% CI: 0.97–1.10). In contrast, the association with inattentiveness remained even after full adjustment (OR: 1.05, 95% CI: 1.12–1.18).

Model 4 includes an interaction term between ADHD and sex and shows that the relationship between ADHD and IPV perpetration differs by sex. Marginal effects analysis (see supplementary information) shows that among individuals without ADHD, men have a slightly higher predicted probability of IPV perpetration than women (9.8% vs. 8.5%). However, among those with ADHD, this pattern reverses: women with ADHD show a substantially higher predicted probability of IPV perpetration than men with ADHD (14.3% vs. 6.9%). Overall, ADHD appears to increase IPV perpetration risk in women, while the association is weaker or even reversed in men. Figure 1.a illustrates this crossover interaction clearly.

**Figure 1.a.**
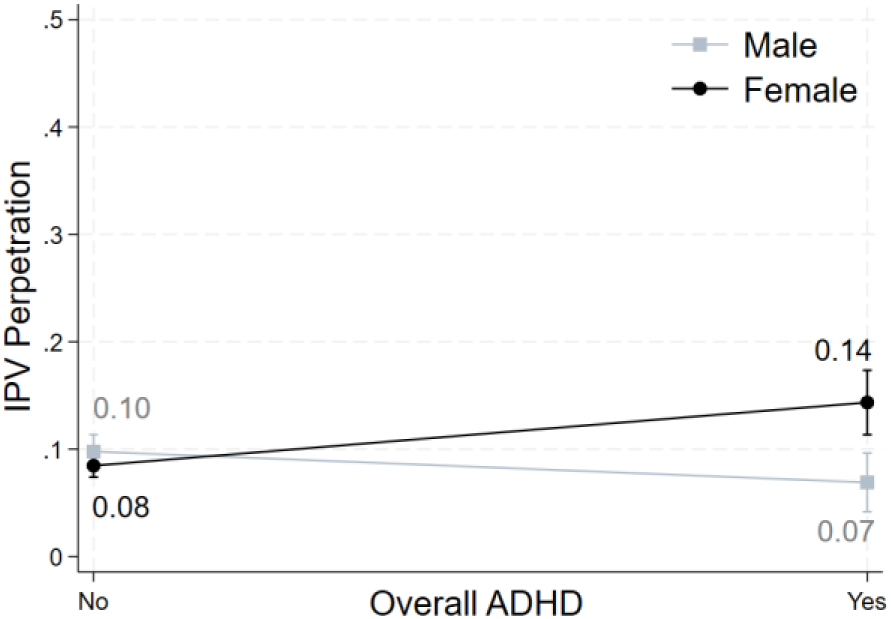
Interaction of Overall ADHD and Sex on IPV Perpetration

Sex interactions are also significant for both ADHD subtypes (Table 3). Marginal effects plots indicate that in the absence of symptoms, males have slightly higher probabilities of IPV perpetration than females (Figure 1.b and Figure 1.c). However, as symptom severity increases, male risk remains relatively stable or declines slightly, while female risk rises steadily, eventually surpassing male risk at moderate-to-high symptom levels. This divergence continues to widen with increasing severity. For inattentiveness, the predicted probability of IPV perpetration reaches 17.5% for women compared to 9.3% for men at the highest score (Figure 1.b). Similarly, at the highest levels of hyperactivity/impulsivity (Figure 1.c and 1.d), the predicted probability is 13.8% for women and 7.3% for men. Notably, at higher severity levels, the 95% CI for the probabilities of IPV perpetration among both males and females are relatively wide for both subtypes.

**Figure 1.b.**
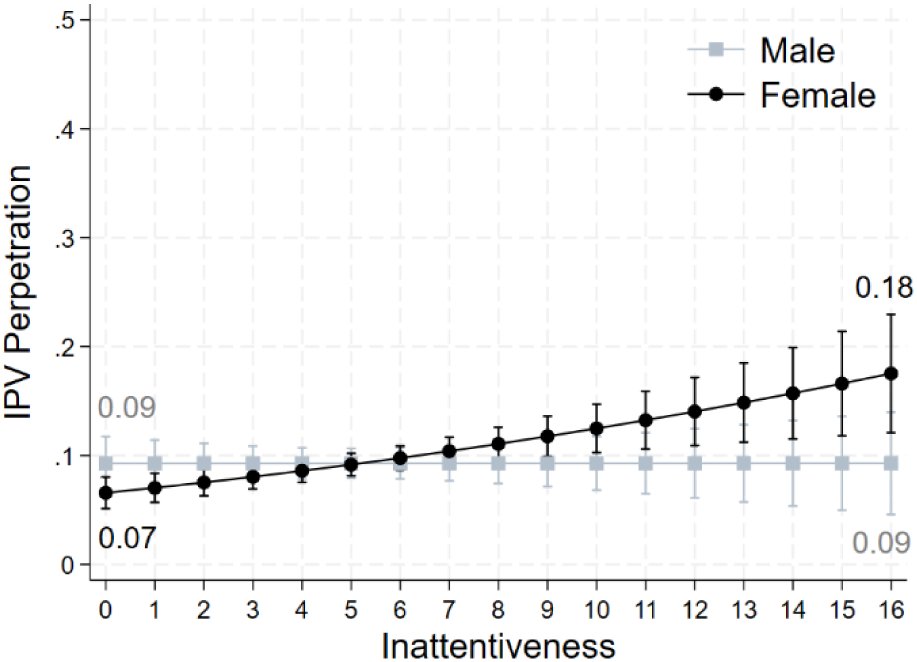
Interaction of Inattentiveness and Sex on IPV Perpetration

**Figure 1.c.**
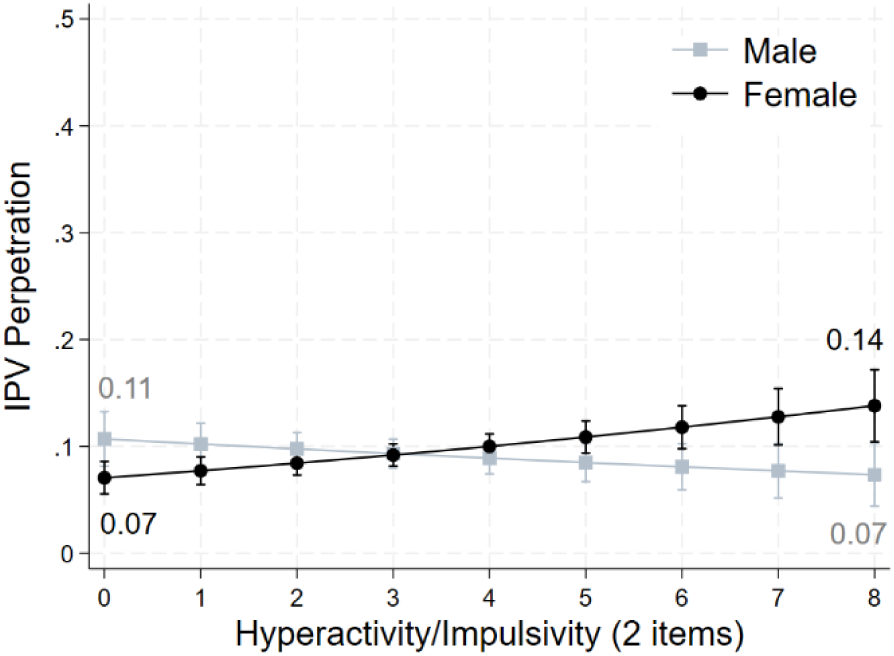
Interaction of Hyperactivity / Impulsivity (2 items) and Sex on IPV Perpetration

**Figure 1.d.**
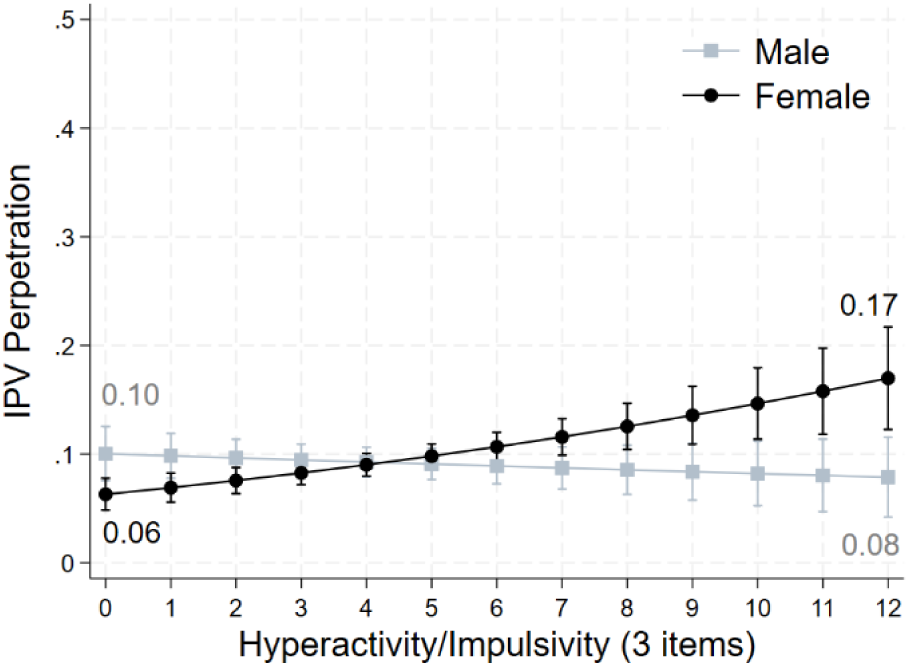
Interaction of Hyperactivity / Impulsivity (3 items) and Sex on IPV Perpetration

The sensitivity analysis using a 3-item hyperactivity/impulsivity measure yielded results broadly consistent with the main analysis (Table 3 and Figure 1.d). The second sensitivity analysis (Model 5), which adjusted only for sociodemographic covariates, also showed aconsistent crossover pattern for the sex-by-ADHD interaction (Supplementary Information). A slight difference was a modest upward trend in IPV perpetration with increasing ADHD severity among men.

## Discussion

This study found that ADHD – both overall and in its inattentive and hyperactive/impulsive subtypes – was positively associated with IPV perpetration after adjusting for sociodemographic factors, consistent with recent systematic reviews (Arrondo et al., 2023; Buitelaar et al., 2020). However, the strength of these associations weakened, and in some cases lost significance, when additional covariates such as IPV victimisation, childhood abuse, non-partner violence perpetration, personality disorder were added sequentially. These findings align with previous research suggesting the confounding or explanatory role of these co-occurring factors (Holmes et al., 2022; Wimberley et al., 2020; Wymbs et al., 2017). Given the cross-sectional nature of the data, the direction or causality of the observed associations, or conclusions regarding confounders or mediators (VanderWeele, 2019), cannot be established.

Among ADHD subtypes, inattentiveness remained associated with IPV perpetration even after full adjustment, while the association with hyperactivity/impulsivity (when measured with three items in the sensitivity analysis) was marginal. This contrasts with Gonzalez et al. (2013), who found an independent effect of hyperactivity/impulsivity but not inattentiveness. While hyperactivity/impulsivity is more commonly linked to aggression (Babinski et al., 2003; Dugre & Potvin, 2022), emerging evidence suggests that inattentiveness may also contribute to violence, potentially through impaired attention switching, emotional recognition, and cognitive control (Bergvall et al., 2001; Gillespie et al., 2015; Romero-Martinez et al., 2019). These deficits may reduce the capacity to interpret social cues and escalate conflict, increasing violence risk.

That the association between ADHD and IPV perpetration differs by sex is a further key finding. Among individuals without ADHD, men reported higher rates of IPV perpetration than women. However, as ADHD symptom severity increased, the risk of IPV perpetration remained relatively stable, or even declined slightly, among men, while it increased gradually among women. At higher symptom levels, women were consequently more likely to report IPV perpetration. These sex-specific patterns were consistent across dimensional assessments and supported by significant interaction effects. This highlights the importance of incorporating sex-disaggregated analyses to elucidate how ADHD and related mental health conditions are associated with interpersonal violence.

Given that ADHD was assessed using a validated screening tool and had comparable prevalence across sexes, diagnostic bias is unlikely to explain these findings. Instead, sex differences may reflect how ADHD symptoms interact with other psychological, relational or contextual risk factors for IPV perpetration. For instance, women with ADHD may be more likely to experience trauma, emotional dysregulation, or relationship conflict, factors which increase the risk of both IPV perpetration and victimisation (Attoe & Climie, 2023). This interpretation aligns with research that has found depression is more strongly associated with IPV perpetration in women than in men (Saunders et al., 2023). While the current study could not assess the motivation or context of IPV perpetration, the finding that IPV victimisation was an important covariate in adjusted models supports the possibility that reciprocal or defensive violence may be relevant for women with ADHD.

It is essential to emphasise that while ADHD was associated with increased risk of IPV perpetration among women, most individuals with ADHD do not engage in violent behaviour. Framing ADHD as a universal risk for violence risks reinforcing stigma and may obscure the complex interplay of factors that shape behavioural outcomes. In addition, wider confidence intervals for the predicted probability of IPV perpetration at higher symptom scores may reflect the smaller number of individuals with severe symptoms.

This study has several methodological strengths. First, it goes beyond estimating average effects by using marginal effects analysis to explore how predicted probabilities of IPV perpetration vary with ADHD symptom severity by sex. This approach offers a more intuitive interpretation than odds ratios and is less sensitive to model specification (Norton & Dowd, 2018). Second, sensitivity analyses were conducted to assess the robustness of findings and methodological assumptions. The comparison between two-item and three-item measures of hyperactivity/impulsivity highlighted the importance of measurement precision in operationalising ADHD subtypes. The model excluding potential mediators enabled a cautious interpretation of sex differences by acknowledging the possibility of causal intermediaries. Third, a broad range of covariates— covering sociodemographic factors, victimisation history, other violent behaviours, and comorbid mental health problems—was included, allowing for a more rigorous assessment of confounding. Finally, IPV perpetration was assessed using a four-item self-completion questionnaire, enhancing disclosure and reducing social desirability bias compared to single-item or interviewer-administered measures (Walby et al., 2014).

Nonetheless, the study has important limitations. First, ADHD medication was excluded from the analysis due to the very small numbers of participants reporting use, precluding investigation of its potentially moderating effects on symptom severity and behavioural outcomes (Buitelaar et al., 2020; Faraone & Buitelaar, 2010; Peterson et al., 2024; Ruiz-Goikoetxea et al., 2018). Second, while overall missing data were limited, previous analyses of the 2014 APMS indicate that missing data on certain variables primarily resulted from non-response to the self-completion section, which was more common among older individuals and those with greater psychological distress (McManus, Gunnell, et al., 2019). Missingness in non-partner violence and personality disorder measures reflects age-based filtering, as SCID-II items were administered only to participants aged 16–64. These patterns may have introduced bias by underrepresenting older and psychologically vulnerable groups. Third, the six-item ASRS is a brief and highly sensitive screening instrument designed to identify individuals likely to have ADHD, but not to provide a formal clinical diagnosis. Additionally, self-reported IPV perpetration is likely to be underreported and may be differentially underreported among men versus women (Emery, 2010). Finally, the cross-sectional design limits the ability to draw causal inferences or determine whether co-occurring conditions function as confounders or mediators.

The study has several implications for service provision, future research, and policy. First, the findings support the development of more tailored, needs-based approaches to IPV perpetration interventions. Traditional programmes, such as Duluth and cognitive-behavioural therapy (CBT) models have shown limited effectiveness, particularly for perpetrators with complex needs (Arias et al., 2013; Babcock et al., 2004). Integrating psychological or psychiatric treatment, including for ADHD and co-occurring conditions, may enhance outcomes for individuals whose IPV perpetration is linked to neurodevelopmental or mental health difficulties.

Additionally, the IPV perpetration variable in this study combined psychological, physical, and sexual violence into a single binary measure. Given known sex differences in the nature and consequences of IPV perpetration behaviours (Hamberger & Larsen, 2015), future studies should disaggregate patterns of IPV behaviours and examine how ADHD interacts with sex across different forms of IPV perpetration.

Longitudinal research is needed to clarify the direction and mechanisms linking ADHD, co-occurring risk factors, and IPV perpetration. Understanding whether variables such as trauma or substance use mediate or confound the relationship between ADHD and IPV is critical for developing preventative interventions. Larger and more diverse samples would further enable examination of potential variations across other intersecting dimensions, including ethnicity and sexuality.

Finally, policy frameworks addressing IPV should account for neurodevelopmental conditions as part of a broader strategy to reduce perpetration. Cross-sector collaboration between IPV services, mental health providers, and neurodevelopmental specialists will be essential to delivering effective, trauma-informed, and equitable responses.

## Conclusion

In conclusion, this study provides novel evidence that ADHD, particularly inattentive symptoms, is associated with increased risk of IPV perpetration, with stronger effects observed among women. While this relationship is partially explained by co-occurring risk factors, the persistence of associations in adjusted models and the clear dose–response relationship with symptom severity suggest that ADHD may contribute to IPV through distinct cognitive and emotional pathways. The findings highlight the need for dimensional assessments of ADHD in both research and clinical practice and underscore the importance of addressing neurodevelopmental conditions within IPV prevention strategies.

## Data Availability

The data analysed in this study are available from the UK Data Service.

http://doi.org/10.5255/UKDA-SN-8203-2

## Notes

### Competing Interest Statement

The authors have declared no competing interest.

### Funding Statement

This study did not receive any funding.

### Author Declarations

Ethics committee of the London School of Hygiene & Tropical Medicine gave ethical approval for this work.

