## Supplementary Information for "Attention-Deficit Hyperactivity Disorder and Intimate Partner Violence Perpetration: Sex Differences and Subtype-Specific Associations in a Population-Based Study"

**Section A: Descriptive analysis of unweighted dataset**

**Table S1.** Unweighted Estimates of Exposures, Outcome, and Covariates by ADHD and IPV Perpetration Status

| Variables | Overall | | | ADHD (n) | | IPV (n) | |
| --- | --- | --- | --- | --- | --- | --- | --- |
|  | n | | % | No | Yes | No | Yes |
| **Exposures** |  | |  |  |  |  |  |
| ADHD | 7341 | |  |  |  | 6212 | 603 |
| No | 6664 | | 90.8 |  |  | 5719 | 477 |
| Yes | 677 | | 9.2 |  |  | 493 | 126 |
| Inattentiveness | 7264 | |  |  |  |  |  |
| *Mean* |  | | 4.1 |  |  | 4.0 | 5.6 |
| *(SD)* |  | | 3.2 |  |  | 3.1 | 3.6 |
| Hyperactivity / Impulsivity(2 items) | 7313 | |  |  |  |  |  |
| *Mean* |  | | 2.4 |  |  | 2.3 | 3.2 |
| *(SD)* |  | | 2.0 |  |  | 2.0 | 2.1 |
| Hyperactivity /Impulsivity (3 items) | 7302 | |  |  |  |  |  |
| *Mean* |  | | 3.3 |  |  | 3.2 | 4.5 |
| *(SD)* |  | | 2.7 |  |  | 2.6 | 2.9 |
| **Outcome** |  | |  |  |  |  |  |
| Lifetime IPV Perpetration | 6825 | |  | 6196 | 619 |  |  |
| No | 6212 | | 91.2 | 5719 | 493 |  |  |
| Yes | 603 | | 8.9 | 477 | 126 |  |  |
| **Sociodemographic Factors** |  | |  |  |  |  |  |
| Ethnicity | 7319 | |  | 6642 | 674 | 6199 | 599 |
| White | 6654 | | 91.0 | 6047 | 606 | 5660 | 554 |
| Black | 194 | | 2.7 | 166 | 28 | 153 | 21 |
| Asian | 325 | | 4.4 | 296 | 28 | 269 | 14 |
| Mixed | 146 | | 2.0 | 133 | 12 | 117 | 10 |
| Sex | 7345 | |  | 6664 | 677 | 6212 | 603 |
| Male | 2936 | | 40.0 | 2669 | 264 | 2504 | 227 |
| Female | 4409 | | 60.0 | 3995 | 413 | 3708 | 376 |
| Age | 7345 | |  |  |  |  |  |
| *Mean* |  | | 52.6 | 53.3 | 45.7 | 52.3 | 47.6 |
| *(SD)* |  | | 18.6 | 18.7 | 16.6 | 18.5 | 15.2 |
| Marital status | 7345 | |  | 6664 | 677 | 6212 | 603 |
| Partnered | 4115 | | 56.0 | 3818 | 295 | 3613 | 282 |
| Non-partnered | 3230 | | 44.0 | 2846 | 382 | 2599 | 321 |
| Employment status | 7,345 | |  | 6664 | 677 | 6,212 | 603 |
| Employed | 3,906 | | 53.2 | 3,595 | 310 | 3,389 | 360 |
| Non-employed | 3,439 | | 46.8 | 3,069 | 367 | 2,823 | 243 |
| **Life Events** |  | |  |  |  |  |  |
| Intimate partner violence victimization | 6,858 | |  | 6,235 | 623 | 6,211 | 603 |
| No | 5,179 | | 75.5 | 4,841 | 338 | 5,000 | 140 |
| Yes | 1,679 | | 24.5 | 1,394 | 285 | 1,211 | 463 |
| Childhood abuse and neglect | 6860 | |  |  |  |  |  |
| *Mean* |  | | 1.9 | 1.8 | 2.8 | 1.7 | 3.3 |
| *(SD)* |  | | 2.4 | 2.3 | 3.0 | 2.9 | 3.1 |
| **Behavioural and Mental Health Disorders** | |  |  |  |  |  |  |
| Perpetration of non-partner violence | 4,900 | |  | 4,344 | 555 | 4,344 | 512 |
| No | 4,619 | | 94.3 | 4,136 | 482 | 4,156 | 426 |
| Yes | 281 | | 5.7 | 208 | 73 | 188 | 86 |
| Personality disorder | 4,917 | |  | 4,360 | 556 | 4,357 | 514 |
| No | 4,679 | | 95.2 | 4,213 | 465 | 4,217 | 419 |
| Yes | 238 | | 4.8 | 147 | 91 | 140 | 95 |
| Substance use | 7,032 | |  | 6,386 | 642 | 6,212 | 603 |
| No | 5,724 | | 81.4 | 5,278 | 442 | 5,129 | 386 |
| Yes | 1,308 | | 18.6 | 1,108 | 200 | 1,083 | 217 |
| **Other** |  | |  |  |  |  |  |
| ADHD medication | 7,342 | |  | 6,662 | 676 | 6,210 | 603 |
| No | 7,339 | | 100.0 | 6,660 | 675 | 6,209 | 602 |
| Yes | 3 | | 0.0 | 2 | 1 | 1 | 1 |

1. Binary and categorical variables are presented as numbers (n) and percentages (%), while continuous variables are shown as means and standard deviations (SD).

2. Ethnicity is shown in original categories to provide descriptive detail; however, a binary variable (White/Minority groups) was used in subsequent analyses to improve model stability.

3. Substance use was originally categorized as: “No”, “Drug dependence / Hazardous alcohol use but no dependency / Alcohol dependence”.

**Section B: Predicted Probability of IPV Perpetration (Main Analyses)**

Note: This section presents the predicted probabilities derived from Model 4. All covariates were included in the model.

**Table S2.** Predicted Probability of IPV Perpetration by Overall ADHD and Sex

|  | Sex | |
| --- | --- | --- |
| Overall ADHD | Male | Female |
| No | 0.0977 | 0.0846 |
| Yes | 0.0690 | 0.1434 |

**Table S3.** Predicted Probability of IPV Perpetration by Inattentiveness and Sex

|  | Sex |  |
| --- | --- | --- |
| Inattentiveness | Male | Female |
| 0 | 0.0928 | 0.0657 |
| 1 | 0.0928 | 0.0704 |
| 2 | 0.0928 | 0.0753 |
| 3 | 0.0928 | 0.0805 |
| 4 | 0.0928 | 0.0859 |
| 5 | 0.0928 | 0.0917 |
| 6 | 0.0928 | 0.0977 |
| 7 | 0.0928 | 0.1040 |
| 8 | 0.0928 | 0.1107 |
| 9 | 0.0928 | 0.1176 |
| 10 | 0.0929 | 0.1249 |
| 11 | 0.0929 | 0.1325 |
| 12 | 0.0929 | 0.1404 |
| 13 | 0.0929 | 0.1486 |
| 14 | 0.0929 | 0.1572 |
| 15 | 0.0929 | 0.1661 |
| 16 | 0.0929 | 0.1753 |

**Table S4.** Predicted Probability of IPV Perpetration by Hyperactivity/Impulsivity (2 items) and Sex

| Hyperactivity/Impulsivity (2 items) | Sex |  |
| --- | --- | --- |
|  | Male | Female |
| 0 | 0.1070 | 0.0706 |
| 1 | 0.1022 | 0.0772 |
| 2 | 0.0977 | 0.0843 |
| 3 | 0.0933 | 0.0919 |
| 4 | 0.0890 | 0.1001 |
| 5 | 0.0849 | 0.1087 |
| 6 | 0.0810 | 0.1180 |
| 7 | 0.0771 | 0.1277 |
| 8 | 0.0735 | 0.1381 |

**Table S5.** Predicted Probability of IPV Perpetration by Hyperactivity/Impulsivity (3 items) and Sex

| Hyperactivity/Impulsivity (2 items) | Sex |  |
| --- | --- | --- |
|  | Male | Female |
| 0 | 0.1004 | 0.0631 |
| 1 | 0.0985 | 0.0691 |
| 2 | 0.0965 | 0.0757 |
| 3 | 0.0946 | 0.0827 |
| 4 | 0.0928 | 0.0902 |
| 5 | 0.0909 | 0.0982 |
| 6 | 0.0891 | 0.1068 |
| 7 | 0.0873 | 0.1159 |
| 8 | 0.0855 | 0.1255 |
| 9 | 0.0838 | 0.1357 |
| 10 | 0.0821 | 0.1465 |
| 11 | 0.0804 | 0.1579 |
| 12 | 0.0788 | 0.1699 |

**Section C: Second Sensitivity Analyses (Model 5)**

Note: This section presents results for the ADHD × Sex interaction, adjusting only for sociodemographic covariates.

**Table S6.** Results of Logistic Regression for ADHD × Sex interaction for IPV Perpetration

|  | Odds ratio | 95% CI (LB) | 95% CI (UB) | p-value |
| --- | --- | --- | --- | --- |
| Overall ADHD | 2.59 | 1.51 | 4.43 | 0.001 |
| Inattentiveness | 1.06 | 1.01 | 1.12 | 0.031 |
| Hyperactivity / impulsivity (2 items) | 1.10 | 1.01 | 1.20 | 0.026 |
| Hyperactivity /impulsivity (3 items) | 1.10 | 1.03 | 1.18 | 0.006 |

**Table S7.** Predicted Probability of IPV Perpetration by Overall ADHD and Sex

|  | Sex | |
| --- | --- | --- |
| Overall ADHD | Male | Female |
| No | 0.0759 | 0.0706 |
| Yes | 0.1137 | 0.2336 |

**Table S8.** Predicted Probability of IPV Perpetration by Inattentiveness and Sex

|  | Sex |  |
| --- | --- | --- |
| Inattentiveness | Male | Female |
| 0 | 0.0504 | 0.0400 |
| 1 | 0.0557 | 0.0469 |
| 2 | 0.0469 | 0.0549 |
| 3 | 0.0678 | 0.0642 |
| 4 | 0.0748 | 0.0749 |
| 5 | 0.0824 | 0.0872 |
| 6 | 0.0907 | 0.1013 |
| 7 | 0.0998 | 0.1173 |
| 8 | 0.1096 | 0.1356 |
| 9 | 0.1203 | 0.1561 |
| 10 | 0.1319 | 0.1791 |
| 11 | 0.1444 | 0.2047 |
| 12 | 0.1579 | 0.2328 |
| 13 | 0.1723 | 0.2635 |
| 14 | 0.1878 | 0.2967 |
| 15 | 0.2043 | 0.3322 |
| 16 | 0.2219 | 0.3696 |

**Table S9.** Predicted Probability of IPV Perpetration by Hyperactivity/Impulsivity (2 items) and Sex

| Hyperactivity/Impulsivity (2 items) | Sex |  |
| --- | --- | --- |
|  | Male | Female |
| 0 | 0.0600 | 0.0477 |
| 1 | 0.0665 | 0.0581 |
| 2 | 0.0736 | 0.0705 |
| 3 | 0.0813 | 0.0853 |
| 4 | 0.0898 | 0.1029 |
| 5 | 0.0991 | 0.1236 |
| 6 | 0.1092 | 0.1477 |
| 7 | 0.1202 | 0.1757 |
| 8 | 0.1322 | 0.2076 |

**Table S10.** Predicted Probability of IPV Perpetration by Hyperactivity/Impulsivity (3 items) and Sex

| Hyperactivity/Impulsivity (2 items) | Sex |  |
| --- | --- | --- |
|  | Male | Female |
| 0 | 0.0562 | 0.0414 |
| 1 | 0.0613 | 0.0495 |
| 2 | 0.0670 | 0.0592 |
| 3 | 0.0731 | 0.0707 |
| 4 | 0.0797 | 0.0842 |
| 5 | 0.0868 | 0.0999 |
| 6 | 0.0946 | 0.1182 |
| 7 | 0.1029 | 0.1393 |
| 8 | 0.1119 | 0.1635 |
| 9 | 0.1216 | 0.1910 |
| 10 | 0.1319 | 0.2218 |
| 11 | 0.1430 | 0.2560 |
| 12 | 0.1549 | 0.2934 |

| **Figure S. a** Interaction of Overall ADHD and Sex on IPV Perpetration 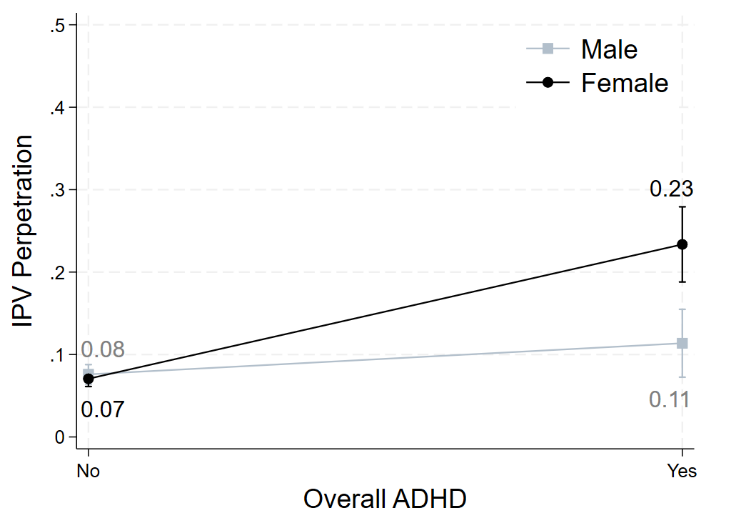 | **Figure S. b** Interaction of Inattentiveness and Sex on IPV Perpetration 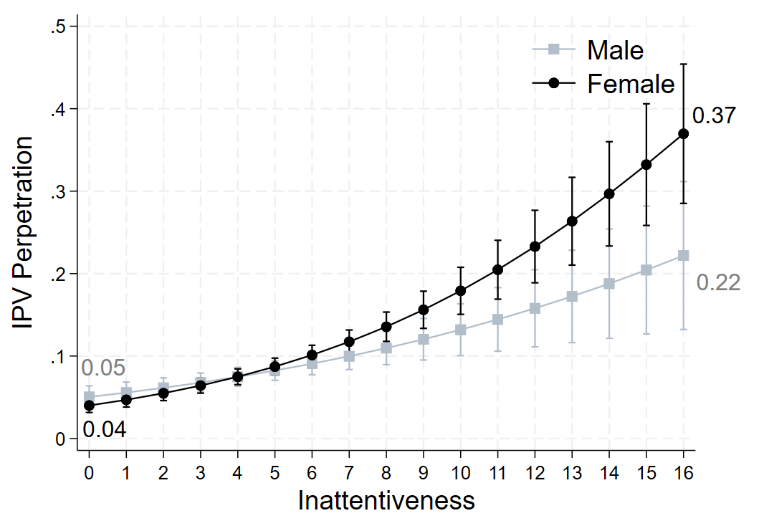 |
| --- | --- |
| **Figure S. c** Interaction of Hyperactivity / Impulsivity (2 items) and Sex on IPV Perpetration 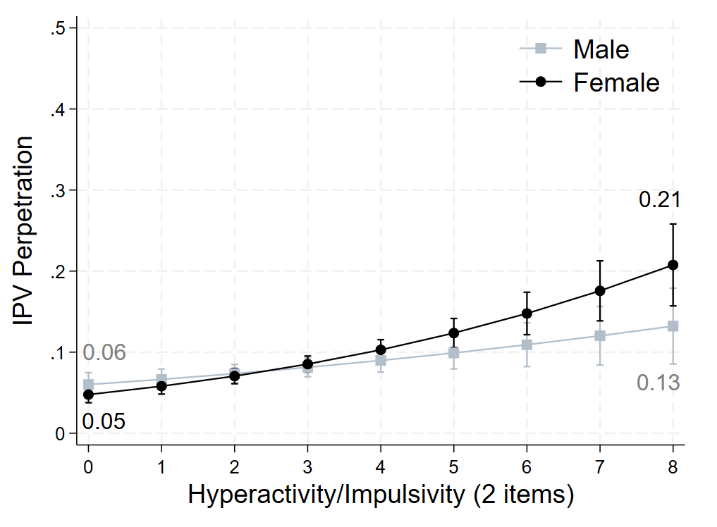 | **Figure S. d** Interaction of Hyperactivity / Impulsivity (3 items) and Sex on IPV Perpetration 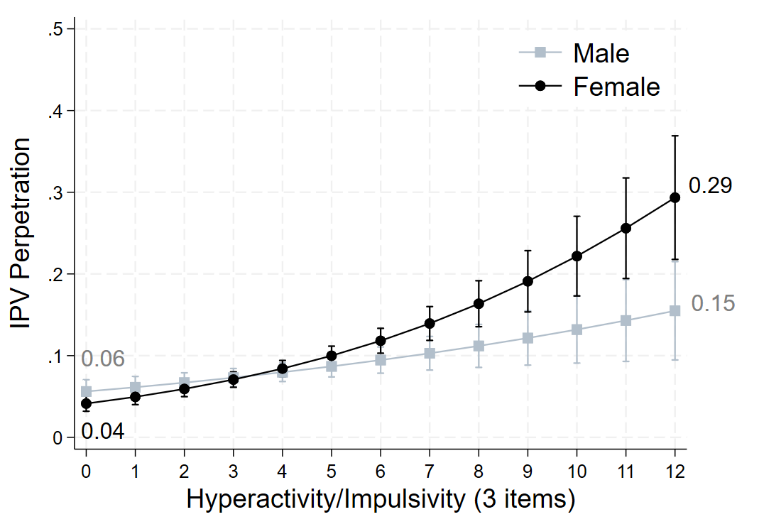 |
